# Predictors of unsuccessful tuberculosis treatment outcomes in Brazil: an analysis of 259,484 patient records

**DOI:** 10.1101/2024.01.05.24300846

**Authors:** Do Kyung Ryuk, Daniele M Pelissari, Kleydson Alves, Patricia Bartholomay Oliveira, Marcia C Castro, Ted Cohen, Mauro Sanchez, Nicolas A Menzies

**Author notes:** Corresponding Author: Nicolas A Menzies, Department of Global Health and Population, Harvard T.H. Chan School of Public Health, 665 Huntington Ave, Boston, MA 02115, USA. **Author contribution statement:** NAM, DR, DMP, KA, PB, MCC, TC, and MS conceptualized the study. NAM and DR designed the analytic strategy. DR performed the analysis. NAM, DR, DMP, KA, PB, MCC, TC, and MS reviewed and interpreted the results. DR drafted the manuscript. NAM, DMP, KA, PB, MCC, TC, and MS reviewed and revised the manuscript.

## Abstract

**Introduction:** Tuberculosis (TB) causes over 1 million deaths annually. Providing effective treatment is a key strategy for reducing TB deaths. In this study, we identified factors associated with unsuccessful treatment outcomes among individuals treated for TB in Brazil.

**Methods:** We obtained data on individuals treated for TB between 2015 and 2018 from Brazil’s National Disease Notification System (SINAN). We excluded patients with a history of prior TB disease or with diagnosed TB drug resistance. We extracted information on patient-level factors potentially associated with successful treatment, including demographic and social factors, comorbid health conditions, health-related behaviors, health system level at which care was provided, use of directly observed therapy (DOT), and clinical examination results. We categorized treatment outcomes as successful (cure, completed) or unsuccessful (death, regimen failure, loss to follow-up). We fit multivariate logistic regression models to identify factors associated with unsuccessful treatment outcome.

**Results:** Among 259,484 individuals treated for drug susceptible TB, 19.7% experienced an unsuccessful treatment outcome (death during treatment 7.8%, regimen failure 0.1%, loss to follow-up 11.9%). The odds of unsuccessful outcome were higher with older age (adjusted odds ratio (aOR) 2.90 [95% confidence interval: 2.62–3.21] for 85-100-year-olds vs. 25-34-year-olds), male sex (aOR 1.28 [1.25– 1.32], vs. female sex), Black race (aOR 1.23 [1.19–1.28], vs. White race), no education (aOR 2.03 [1.91– 2.17], vs. complete high school education), HIV infection (aOR 2.72 [2.63–2.81], vs. no HIV infection), illicit drug use (aOR 1.95 [1.88–2.01], vs. no illicit drug use), alcohol consumption (aOR 1.46 [1.41– 1.50], vs. no alcohol consumption), smoking (aOR 1.20 [1.16–1.23], vs. non-smoking), homelessness (aOR 3.12 [2.95–3.31], vs. no homelessness), and immigrant status (aOR 1.27 [1.11–1.45], vs. non-immigrants). Treatment was more likely to be unsuccessful for individuals treated in tertiary care (aOR 2.20 [2.14–2.27], vs. primary care), and for patients not receiving DOT (aOR 2.35 [2.29–2.41], vs. receiving DOT).

**Conclusion:** The risk of unsuccessful TB treatment varied systematically according to individual and service-related factors. Concentrating clinical attention on individuals with a high risk of poor treatment outcomes could improve the overall effectiveness of TB treatment in Brazil.

## Introduction

Tuberculosis (TB) is a major cause of infectious disease morbidity and mortality globally [1]. In 2022, 1.3 million individuals are estimated to have died with TB, representing one out of every seven individuals that develop TB. Providing early diagnosis and effective treatment is a key strategy for reducing TB deaths. In the absence of drug resistance, TB is treated using a standardized 6-month course of rifampin, isoniazid, ethambutol, and pyrazinamide [3].

Following standardized reporting guidelines, the outcomes of TB treatment are categorized as one of five mutually-exclusive categories: cured, treatment completed (regimen successfully completed but without smear reversion from positive to negative), lost to follow-up, (4) dead, (5) and treatment failed [4]. Treatment failure or loss to follow-up can result in a longer duration of disease, elevated mortality risks, and the possibility of acquired drug resistance [1]. To avoid these negative outcomes, it is important to implement effective patient-centric strategies to increase the fraction of patients achieving successful TB treatment outcomes.

Brazil is one of thirty high TB burden countries identified by the World Health Organization (WHO) [1]. In Brazil, TB diagnosis and treatment is provided through the universal healthcare system, Sistema Único de Saúde (SUS), under which TB incidence and mortality have decreased over time [5]. However, this decline stalled following an economic crisis in 2014 [6] and TB mortality was estimated as 3.3 per 100,000 in 2019 [7, 8]. During the COVID-19 pandemic, the national TB program reported an increase in loss to follow-up, which in 2020 rose to 12% of treated individuals, and has also experienced decreasing rates of participation in directly observed therapy (DOT) [5]. In addition, the proportion of patients who are recorded as being cured has steadily decreased, from 73% in 2018 to 65% in 2020 [5].

Understanding how different factors are associated with TB treatment outcomes can suggest approaches to improving care, and help identify patients with the greatest risks of experiencing unsuccessful outcomes. In this study, we assessed potential risk factors for unsuccessful TB treatment outcomes under routine clinical conditions in Brazil’s national TB treatment program. Using national disease registry data, we analyzed treatment outcomes for individuals initiating TB treatment between 2015 and 2018. We estimated how treatment outcomes varied by demographic and socio-economic factors, the presence of co-morbidities, health-related behaviors, and features of service provision, as well as how outcomes varied across Brazilian states.

## Method

### Data sources

We obtained data on all individuals treated for TB disease between 2015 and 2018 (n=356,119) from Brazil’s National Disease Notification Information System (SINAN: Sistema de Informação de Agravos de Notificação). These data include individuals diagnosed with TB, including pulmonary and extrapulmonary disease, in all 26 Brazilian states and the Federal District (Brasília).

We excluded patients previously diagnosed with TB (n=68,519, 19.2%), patients diagnosed with resistance to rifampicin (n=2,584, 0.7%), patients who had a change in regimen due to adverse event or identified drug-resistance (n=2,019, 0.6%), patients transferred to a different provider during therapy (n=20,306, 5.7%), patients diagnosed with TB post-mortem (n=2,695, 0.8%), patients with a missing value for treatment outcome (n=10,786, 3.0%), and patients with illogical values for exposure variables, such as miscategorized age (n=56, <0.1%) [9]. For each individual included in the study cohort we extracted information on patient-level factors potentially associated with TB treatment outcomes (Table 1). These include socio-demographics (sex, age, education, self-declared race), vulnerability status (incarcerated, homelessness, immigrants), other health conditions (HIV, diabetes), health-related behaviors (illicit drug use, alcohol consumption, current smoking), type of TB disease (pulmonary, extrapulmonary, or both), aspects of clinical care (participation in DOT, pre-treatment diagnostic test results (bacteriological diagnosis, chest x-ray)), and the health system level at which treatment was provided (obtained through linkage between SINAN and the National Registry of Health Establishment (CNES)). We also recorded the state where each individual received treatment.

**Table 1.** Definitions of outcome and exposure variables. DOT = directly observed therapy. HIV = human immunodeficiency virus. “Other” category for variable HIV refers to patients who have their test in progress, or patients who did not test. “Other” category for variables such as diabetes, illicit use of drugs, alcohol, smoking, DOT, patients in vulnerable circumstances (incarcerated, homeless, immigrant) includes patients who did not respond to the question. “Other” category for the level of health service variable includes laboratory centers or private clinics.

| Variable | Variable definition |
| --- | --- |
| ***Outcome variables*** |  |
| Unsuccessful treatment outcome<br>(*main analysis*) | Yes (includes loss to follow-up, death, failure), No (includes completion, cure) |
| Categorical treatment outcome<br>(*secondary analysis*) | Death, loss to follow-up, success (includes completion, cure) |
| ***Exposure variables*** |  |
| Age group | Categorized as 0-4, 5-14, 15-24, 25-34, 35-44, 45-54, 55-64, 65-74, 75-84, or 85-100 years of age at diagnosis. |
| Sex | Categorized as male or female |
| Education level | Categorized as no education, incomplete/complete 1-4<sup>th</sup> grade, complete 5-8<sup>th</sup> grade, complete high school education, any higher education, or other |
| Race | Categorized as White, Black, Yellow, Mixed, Indigenous, or other |
| HIV | Categorized as yes, no, or other |
| Diabetes | Categorized as yes, no, or other |
| Illicit drug use | Categorized as yes, no, or other |
| Alcohol use | Categorized as yes, no, or other |
| Smoking | Categorized as yes, no, or other |
| Incarcerated | Categorized as yes, no, or other |
| Homeless | Categorized as yes, no, or other |
| Immigrant | Categorized as yes, no, or other |
| Level of health service | Categorized as primary, secondary, tertiary, or other |
| Received DOT | Categorized as yes, no, or other |
| Bacteriological test result | Categorized as positive, negative, or not determined |
| Chest x-ray result | Categorized as presumed with TB, normal, or not performed |
| Type of TB | Categorized as pulmonary, extrapulmonary or both |

### Outcome definition

In SINAN, individuals treated for TB can have a treatment outcome recorded as ‘treatment success’ (completion of treatment with two or more successive negative sputum smear microscopy results), representing the sum of ‘cured’ and ‘treatment completed’ treatment outcome categories defined by WHO [4]. Individuals recorded with ‘death on treatment’ (defined as death from TB or other cause during TB treatment), ‘regimen failure’ (defined as having positive sputum smear or culture in the 4th month or two consecutive months after the 4th month of treatment initiation) or ‘loss to follow-up’(defined as the patient not attending the treatment facilities for 30 days or more once treatment has started) were coded as having an unsuccessful treatment outcome [10].

For the main analysis we analyzed a binary outcome indicating whether the individual experienced an unsuccessful treatment outcome. As a secondary analysis we analyzed a categorical outcome with three levels (treatment success, loss to follow-up, and death) to allow for different predictors of loss to follow-up and death. For this secondary analysis, we did not consider the outcome of treatment failure, given the small number of individuals in this group.

### Statistical analysis

We fitted univariate and multivariate logistic regression models to identify factors associated with unsuccessful treatment outcomes, considering each exposure variable as well as state of residence. For most variables we selected the category with the highest number of observations as the reference group. For race and education level, we selected ‘White’ and ‘completed high-school education’ (respectively) as the reference categories, representing population groups historically associated with better TB outcomes, such that the results describe the excess risks faces by other populations. Results are reported as odds ratios. For the secondary analysis of categorical treatment outcome (success, loss to follow-up, death), we fitted multinomial logistic regression models to estimate the factors associated with specific treatment outcomes. As a sensitivity analysis, we refit separate regression models for the binary treatment outcome to data for each calendar year.

We conducted additional analyses to estimate the importance of each exposure variable in explaining treatment outcomes within the study cohort. To do so, we refit the main analysis regression model (for the binary treatment outcome) excluding each covariate one at a time, and estimated Akaike Information Criterion (AIC) for each of these models. We calculated the difference between these values and the AIC estimated for the full model including all the covariates, reporting these difference measures as an indicator of variable importance. We calculated confidence intervals for these results using a bootstrap approach with 100 replicates. All analyses were conducted in R [10].

## Results

Table 2 describes the distribution of individuals across levels of each exposure variable. Among 259,484 individuals included in the study cohort, 19.7% (n=51,160) experienced an unsuccessful treatment outcome (death on treatment 7.8%, regimen failure 0.1%, loss to follow-up 11.9%).

**Table 2.** Baseline demographic information of the study population. DOT = directly observed therapy. HIV = human immunodeficiency virus.

| Variables (Reference) | Number<br>(percentage) | Variables (Reference) | Number<br>(percentage) |
| --- | --- | --- | --- |
| **Age group (25-34)** | 64,688 (22.7%) | **Alcohol (no)** | 202,673 (78.1%) |
| 0-4 | 3,018 (1.2%) | Yes | 41,723 (16.1%) |
| 5-14 | 4,841 (1.9%) | Other | 15,088 (5.8%) |
| 15-24 | 49,267 (19.0%) | **Drug (no)** | 209,783 (80.8%) |
| 35-44 | 47,682 (18.4%) | Yes | 30,380 (11.7%) |
| 45-54 | 39,913 (15.4%) | Other | 19,321 (7.4%) |
| 55-64 | 30,697 (11.8%) | **Incarcerated (no)** | 218,693 (84.3%) |
| 65-74 | 15,884 (6.1%) | Yes | 24,882 (9.6%) |
| 75-84 | 7,363 (2.8%) | Other | 15,909 (6.1%) |
| 85+ | 1,994 (0.8%) | **Homeless (no)** | 236,215 (91.0%) |
| **Sex (Male)** | 177,330 (68.3%) | Yes | 6,172 (2.4%) |
| Female | 82,160 (31.7%) | Other | 17,097 (6.6%) |
| **Race (White)** | 82,426 (31.8%) | **Immigrants (no)** | 236,979 (91.3%) |
| Black | 31,404 (31.8%) | Yes | 1,543 (0.6%) |
| Yellow | 1,825 (12.1%) | Other | 20,962 (8.1%) |
| Mixed | 122,125 (47.1%) | **Health unit (primary care)** | 140,807 (54.3%) |
| Indigenous | 2,909 (1.1%) | Secondary care | 74,173 (28.6%) |
| Other | 18,795 (7.2%) | Tertiary care | 35,854 (13.8%) |
| **Education (complete high school)** | 24,137 (9.3%) | Other | 8,650 (3.3%) |
| No education | 11,179 (4.3%) | **DOT (yes)** | 99,353 (38.3%) |
| Incomplete 1-4th grade | 29,434 (11.3%) | No | 97,422 (37.5%) |
| Complete 1-4<sup>th</sup> grade | 60,779 (23.4%) | Other | 62,709 (24.2%) |
| Complete 5-8<sup>th</sup> grade | 47,932 (18.5%) | **Bacteriological test (positive)** | 169,512 (65.3%) |
| Any higher education | 16,596 (6.4%) | Negative | 44,195 (17.0%) |
| Other | 69,427 (26.8%) | Not determined | 45,777 (17.6%) |
| **Diabetes (no)** | 223,818 (86.3%) | **Chest X-ray (suggestive with TB)** | 183,784 (70.8%) |
| Yes | 19,937 (7.7%) | Normal | 16,977 (6.5%) |
| Other | 15,729 (6.1%) | Not performed | 58,723 (22.6%) |
| **HIV (negative)** | 191,119 (73.7%) | **Type of TB (pulmonary)** | 217,486 (83.8%) |
| Yes | 23,328 (9.0%) | Extrapulmonary | 7,500 (2.9%) |
| Other | 45,037 (17.4%) | Both | 34,498 (13.3%) |
| **Smoking (no)** | 188,028 (72.5%) |  |  |
| Yes | 53,390 (20.6%) |  |  |
| Other | 18,066 (7.0%) |  |  |

### Odds ratios for unsuccessful treatment outcome

Unadjusted and adjusted odds ratios for unsuccessful treatment outcome for each exposure variable are reported in Table 3, based on the results of univariate and multivariate regression models, respectively. Significant differences in the odds of an unsuccessful treatment outcome were estimated for several exposure variables. We estimated elevated risks of unsuccessful treatment outcome (adjusted odds ratios (aORs) >1.0) for variables describing age >65 years (versus age 25-34), Black race (versus White race), educational level less than complete high school education (versus complete high school education), HIV-positive or HIV unknown status (versus HIV-negative), smoking (versus non-smoking), alcohol consumption (versus no alcohol consumption), illicit drug usage (versus no illicit drug use), homelessness (versus no homelessness), immigrant status (versus non-immigrants), treatment provision in secondary or tertiary care (versus primary care), not enrolled in DOT therapy (versus DOT), bacteriological test negative or not determined (versus individuals with a positive bacteriological test result), and chest x-ray not performed (versus x-ray suggestive of TB). Age < 15 years (versus age 25-34), female sex (versus male sex), education above high school level (versus complete high school education) and diabetes (versus no diabetes) were associated with lower risks of unsuccessful treatment outcome.

**Table 3.** Raw and adjusted odds ratio for unsuccessful treatment outcome for each exposure variable, 2015-2018. DOT = directly observed therapy. HIV = human immunodeficiency virus. CI = confidence interval. Raw odds ratios estimated from regression models including each exposure variable individually. Adjusted odds ratios estimated from a regression model including all exposure variables.

| Variables (reference category) | Univariate odds ratio (95% CI) | Adjusted odds ratio (95% CI) |
| --- | --- | --- |
| **Age (25-34)** |  |  |
| 0-4 | 0.65 (0.59, 0.73) | 0.50 (0.45, 0.56) |
| 5-14 | 0.41 (0.37, 0.46) | 0.39 (0.35, 0.43) |
| 15-24 | 0.87 (0.84, 0.90) | 1.04 (1.01, 1.08) |
| 35-44 | 1.13 (1.10, 1.17) | 0.93 (0.90, 0.97) |
| 45-54 | 1.04 (1.01, 1.07) | 0.91 (0.88, 0.94) |
| 55-64 | 1.00 (0.97, 1.04) | 0.98 (0.94, 1.02) |
| 65-74 | 1.24 (1.19, 1.29) | 1.25 (1.19, 1.31) |
| 75-84 | 1.77 (1.68, 1.87) | 1.90 (1.79, 2.02) |
| 85+ | 2.93 (2.68, 3.21) | 2.90 (2.62, 3.21) |
| **Sex (Male)** |  |  |
| Female | 0.70 (0.68, 0.71) | 0.78 (0.76, 0.80) |
| **Race (White)** |  |  |
| Black | 1.42 (1.37, 1.47) | 1.23 (1.19, 1.28) |
| Yellow | 1.01 (0.90, 1.15) | 0.99 (0.87, 1.12) |
| Mixed | 1.19 (1.17, 1.22) | 1.14 (1.11, 1.12) |
| Indigenous | 0.81 (0.73, 0.90) | 1.05 (0.94, 1.17) |
| Other | 1.36 (1.30, 1.41) | 1.02 (0.98, 1.07) |
| **Education (complete high school)** |  |  |
| No education | 2.04 (1.93, 2.16) | 2.03 (1.91, 2.17) |
| Incomplete 1-4<sup>th</sup> grade | 1.77 (1.69, 1.86) | 1.80 (1.71, 1.90) |
| Complete 1-4<sup>th</sup> grade | 1.73 (1.65, 1.80) | 1.83 (1.74, 1.91) |
| Complete 5-8<sup>th</sup> grade | 1.32 (1.26, 1.38) | 1.46 (1.39, 1.53) |
| Any higher education | 0.72 (0.67, 0.76) | 0.84 (0.79, 0.90) |
| Other | 2.15 (2.06, 2.24) | 1.94 (1.85, 2.03) |
| **Diabetes (no)** |  |  |
| Yes | 0.93 (0.89, 0.96) | 0.91 (0.87, 0.94) |
| Other | 1.60 (1.54, 1.66) | 0.90 (0.85, 0.97) |
| **HIV (no)** |  |  |
| Yes | 3.87 (3.76, 3.98) | 2.72 (2.63, 2.81) |
| Other | 2.24 (2.19, 2.30) | 1.83 (1.78, 1.88) |
| **Smoking (no)** |  |  |
| Yes | 1.67 (1.63, 1.70) | 1.20 (1.16, 1.23) |
| Other | 1.81 (1.75, 1.87) | 1.10 (1.02, 1.18) |
| **Alcohol (no)** |  |  |
| Yes | 2.18 (2.13, 2.23) | 1.46 (1.41, 1.50) |
| Other | 1.85 (1.78, 1.92) | 1.09 (1.01, 1.17) |
| **Illicit drug use (no)** |  |  |
| Yes | 2.48 (2.42, 2.55) | 1.95 (1.88, 2.01) |
| Other | 1.81 (1.75, 1.87) | 1.21 (1.13, 1.30) |
| **Incarcerated (no)** |  |  |
| Yes | 0.60 (0.58, 0.62) | 0.52 (0.49, 0.54) |
| Other | 1.26 (1.21, 1.30) | 1.18 (1.05, 1.32) |
| **Homeless (no)** |  |  |
| Yes | 5.26 (4.99, 5.53) | 3.12 (2.95, 3.31) |
| Other | 1.33 (1.28, 1.38) | 0.87 (0.77, 0.99) |
| **Immigrants (no)** |  |  |
| Yes | 1.31 (1.17, 1.48) | 1.27 (1.11, 1.45) |
| Other | 1.22 (1.18, 1.26) | 0.92 (0.85, 1.00) |
| **Health unit (primary care)** |  |  |
| Secondary care | 1.59 (1.55, 1.62) | 1.20 (1.17, 1.24) |
| Tertiary care | 2.94 (2.87, 3.02) | 2.20 (2.14, 2.27) |

| **Variables (reference category)** | **Univariate odds ratio (95% CI)** | **Adjusted odds ratio (95% CI)** |
| --- | --- | --- |
| Other | 1.22 (1.15, 1.29) | 1.02 (0.95, 1.08) |
| **DOT (yes)** |  |  |
| No | 2.42 (2.36, 2.48) | 2.35 (2.29, 2.41) |
| Other | 3.74 (3.65, 3.84) | 3.13 (3.04, 3.22) |
| **Bacteriological test (positive)** |  |  |
| Negative | 1.15 (1.12, 1.18) | 1.14 (1.11, 1.18) |
| Not determined | 1.29 (1.26, 1.32) | 1.33 (1.29, 1.38) |
| **Chest X-ray (suggestive of TB)** |  |  |
| Normal | 0.91 (0.87, 0.94) | 0.95 (0.91, 1.00) |
| Not performed | 0.88 (0.86, 0.90) | 1.03 (1.00, 1.06) |
| **Type of TB (pulmonary)** |  |  |
| Extrapulmonary | 1.71 (1.63, 1.80) | 1.03 (0.98, 1.09) |
| Both | 0.88 (0.86, 0.91) | 0.72 (0.69, 0.75) |

For most exposure variables univariate ORs were similar to the results of the multivariate analysis. However, univariate ORs for HIV, smoking, alcohol consumption, illicit drug use, and homelessness were elevated compared to adjusted ORs, consistent with clustering of these risk factors within a subset of patients experiencing worse treatment outcomes. In sensitivity analyses we refit separate regression models to the data for each calendar year (Supplementary Table 2). These results were similar to those estimated in the main analysis.

### State-level differences in treatment outcome

At the state level, the univariate model described a highest odds of unsuccessful treatment outcome in the state of Rio Grande do Sul (OR=1.78, 95% CI: 1.71-1.85), and the lowest odds of unsuccessful treatment outcome in the state of Acre (OR=0.38, 95% CI: 0.31-0.47), both compared to the state of São Paulo (Figure 1). Adjusted odds ratios (controlling for all other exposure variables) described the highest odds of unsuccessful treatment outcome in the state of Roraima (aOR=1.67, 95% CI: 1.35-2.06), and the lowest odds in the state of Acre (aOR=0.58, 95% CI: 0.47-0.71).

**Figure 1.**
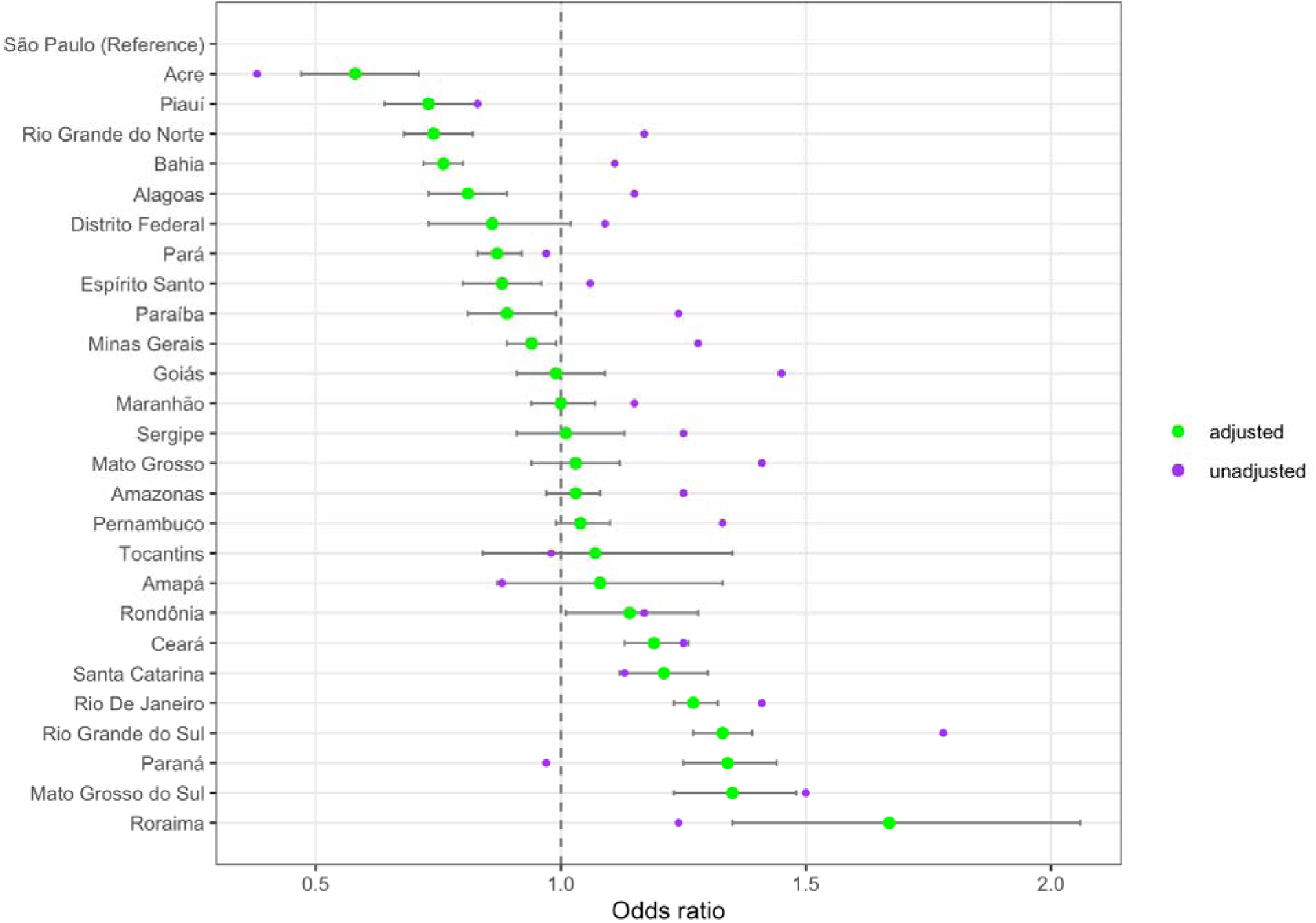
Unadjusted and adjusted odds ratios of unsuccessful treatment outcome for each state, 2015-2018. Unadjusted odds ratios estimated from regression models including each exposure variable individually. Adjusted odds ratios estimated from a regression model including all exposure variables. Horizontal bars represent 95% confidence intervals for adjusted odds ratios.

### Importance of individual exposure variables and sensitivity analysis

Table 4 presents results describing the relative importance of each exposure variable. Based on these results, treatment via DOT, HIV status, healthcare level of the treatment provider, education level, and age group were the most important variables in terms of explaining the variation in treatment outcomes within the study cohort.

**Table 4.** Importance of each exposure variable for explaining cohort treatment outcomes. DOT = directly observed therapy. HIV = human immunodeficiency virus. Variable importance calculated from the difference in AIC for models excluding each exposure variable as compared to the full regression model for the binary treatment outcome (AIC = 222461). Greater values indicate greater importance for a given exposure variable.

| **Exposure variable** | **Variable importance\***<br>**(95% CI)** |
| --- | --- |
| Chest x-ray result | 5 (-3, 19) |
| Diabetes | 11 (1, 34) |
| Immigrant status | 16 (2, 32) |
| Smoking status | 136 (89, 184) |
| Race | 214 (160, 261) |
| Bacteriological test result | 329 (272, 414) |
| Type of TB | 343 (280, 423) |
| Sex | 363 (315, 448) |
| Alcohol use | 517 (427, 626) |
| Incarceration | 912 (808, 1,021) |
| Illicit drug use | 1,473 (1,326, 1,609) |
| Homeless | 1,523 (1,371, 1,694) |
| Age group | 1,661 (1,517, 1,839) |
| Education level | 1,731 (1,562, 1,885) |
| Health care level of treatment provider | 2,506 (2,338, 2,702) |
| HIV status | 4,906 (4,595, 5,148) |
| DOT received | 7,294 (6,904, 7,599) |

### Risk ratios for categorical outcome

Supplementary Table 3 presents results for the categorical treatment outcome (success, death, loss to follow-up) estimated via multinomial logistic regression analysis. For several variables, the factors associated with loss to follow-up differ from those associated with death on treatment. In particular, while older age groups experienced higher mortality rates (versus 15-24-year-olds), risks of loss to follow-up were lower. Conversely, adjusted risk ratios (aRRs) describing elevated risks of loss to follow-up for Black, Yellow or Mixed race, illicit drug use, homelessness, and immigrant status were all comparatively greater than the aRRs describing risks of death. For location of care, care provided in secondary and tertiary sites was strongly associated with death while these relationships were weaker for loss to follow-up. Treatment provided via DOT was associated with lower risks of both death and loss to follow-up, with this relationship being stronger for loss to follow-up.

## Discussion

In this study we examined the relationship between treatment outcomes and individual demographics, pre-existing conditions, health-related behaviors, membership of special populations, clinical examination results, and features of health services among individuals treated for TB in Brazil between 2015 and 2018. These analyses revealed elevated risks of unsuccessful TB treatment outcomes associated with a range of demographic, clinical and behavioral factors. Unless addressed in some way, these excess risks will make it difficult for Brazil to meet the End TB Strategy target of 90% of individuals achieving successful TB treatment outcomes [8].

In terms of socio-demographic and behavioral factors, the strongest relationships with unsuccessful treatment outcomes were estimated for old age, no education or limited education, HIV infection, illicit drug use, and homelessness. Elevated mortality on treatment was found to be the primary cause of poor treatment outcomes for individuals with HIV and old age, while elevated loss to follow-up was the most important factors for homeless individuals and those with illicit drug use. Both factors were found to be important for individuals with no education or limited education. These findings are consistent with previous systematic reviews and meta-analyses [11–13], and point to the greater challenges of achieving successful treatment outcomes for medically fragile individuals, and for individuals with vulnerable circumstances or health behaviors that make it more difficult to complete the extended treatment regimens required for TB disease. Treatment completion was found to be higher among incarcerated patients, consistent with earlier studies [12, 16, 17]. However, TB treatment completion among incarcerated individuals may be eroded when patients are transferred between facilities or released during treatment, as coordination of care is often challenging [17].

In terms of clinical factors, our results revealed a strong relationship between the risk of unsuccessful treatment outcomes and enrollment in DOT. Individuals who enrolled in DOT were substantially more likely to experience a successful treatment outcome, and DOT treatment was associated with lower risks of both loss to follow-up and death on treatment. It is possible these relationships are not consistent across Brazil, as the approach to providing DOT differ at the state level [14, 15]. The greater success rates experienced with DOT treatment must be interpreted carefully, as it will reflect both the impact of DOT through supporting better treatment adherence and completion (the causal effect), as well as differences in treatment outcomes resulting from differences in the characteristics of patients enrolled versus not enrolled in DOT (the non-casual effect). However, the large magnitude of this effect demonstrates the importance of DOT enrollment in understanding TB treatment outcomes in this setting. This is also shown in the results for the variable importance analysis, which found DOT to be the most important single factor for predicting treatment outcomes in this study population. As traditional DOT requires patients to consume drugs on-site multiple times per week, this can cause challenges for some patients (particularly those in vulnerable situations) and limit the proportion of patients enrolled in DOT. To address this challenge, the Brazilian health system is considering alternative DOT modalities that do not require in-person attendance (e.g, video-based DOT). If successful, these new DOT modalities could raise DOT enrollment and enhance treatment adherence (particularly in groups with currently low rates of treatment success), as well as giving patients greater autonomy over when and where they take their medication.

The health system level at which TB treatment is provided was also found to be strongly related to the risk of unsuccessful treatment. Controlling for other factors, patients treated in primary facilities were less likely to experience an unsuccessful treatment outcome compared to those treated in secondary or tertiary facilities. As higher-level clinical facilities typically treat individuals with more complex disease cases, it is likely the results for this variable reflect differences in case-mix between health system levels, not sufficiently captured by the other variables included in the analysis [9]. However, the high levels of unsuccessful outcome experienced by patients at higher-level facilities indicates the potential for greater absolute improvements in outcomes in these settings.

Several previous studies conducted in low- and middle-income countries have focused on specific factors associated with the TB treatment outcome, such as HIV co-infection, TB drug resistance, and social vulnerability [18, 19, 20]. Our study adds to this literature by using national registry data to identify the patient subgroups that are at greater risk of poor treatment outcomes. However, this study has several limitations. Most importantly, the relationships estimated in this analysis represent statistical associations rather than causal relationships. As a consequence, while the results can be used to describe patient subgroups that are at high risk of poor outcomes—and that would potentially benefit from greater clinical attention—they do not describe the improvements in outcomes that could be achieved by changes in patient care, such as by devolving more TB care to the primary facilities or increasing DOT enrollment. Second, the outcome examined (treatment success) has limitations as an indicator of treatment effectiveness. In particular, some individuals coded as treatment success will not have achieved sterilizing cure and will go on to relapse in the years following treatment. While these relapse cases may be identified in research cohorts, they are not linked to the original treatment episode in the disease registry data. Third, we did not investigate interactions between exposure variables, or how the estimated relationships varied across states. Given the differences in TB care and populations characteristics across Brazil, it is possible such variation exists.

## Conclusion

The fraction of patients experiencing unsuccessful TB treatment outcomes varies systematically as a function of socio-demographic factors, co-morbidities, health-related behaviors, clinical presentation, and features of clinical of care. Focusing clinical attention on patients with these risk factors could improve overall program performance and reduce disparities in treatment outcomes between population groups. Future research is needed to develop scalable treatment modalities that support regimen adherence and treatment completion, particularly among population groups with life circumstances that make this challenging.

## Data Availability

All data used in this study are publicly available. TB case and mortality data can be accessed at: https://datasus.saude.gov.br/. Demographic data can be accessed at: https:// www.ibge.gov.br/. Level of health care can be accessed at: https://dados.gov.br/dados/conjuntos-dados/cnes-cadastro-nacional-de-estabelecimentos-de-saude .

https://www.ibge.gov.br/

https://datasus.saude.gov.br/

## SUPPLEMENTARY TABLE

**Table S1.**
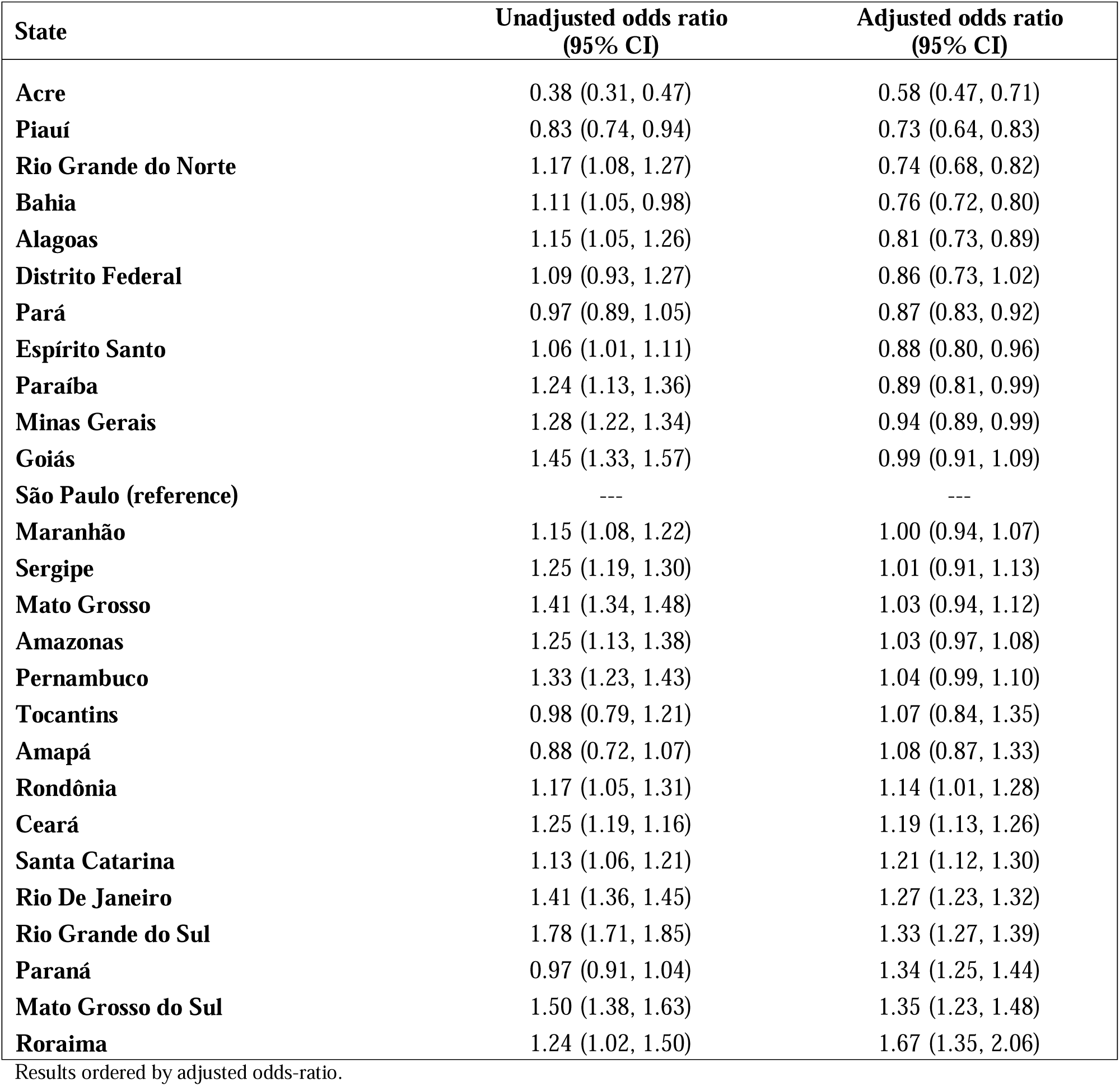
Unadjusted and adjusted odds ratios for unsuccessful treatment outcome at state level, 2015-2018.

| State | Unadjusted odds ratio<br>(95% CI) | Adjusted odds ratio<br>(95% CI) |
| --- | --- | --- |
| **Acre** | 0.38 (0.31, 0.47) | 0.58 (0.47, 0.71) |
| **Piauí** | 0.83 (0.74, 0.94) | 0.73 (0.64, 0.83) |
| **Rio Grande do Norte** | 1.17 (1.08, 1.27) | 0.74 (0.68, 0.82) |
| **Bahia** | 1.11 (1.05, 0.98) | 0.76 (0.72, 0.80) |
| **Alagoas** | 1.15 (1.05, 1.26) | 0.81 (0.73, 0.89) |
| **Distrito Federal** | 1.09 (0.93, 1.27) | 0.86 (0.73, 1.02) |
| **Pará** | 0.97 (0.89, 1.05) | 0.87 (0.83, 0.92) |
| **Espírito Santo** | 1.06 (1.01, 1.11) | 0.88 (0.80, 0.96) |
| **Paraíba** | 1.24 (1.13, 1.36) | 0.89 (0.81, 0.99) |
| **Minas Gerais** | 1.28 (1.22, 1.34) | 0.94 (0.89, 0.99) |
| **Goiás** | 1.45 (1.33, 1.57) | 0.99 (0.91, 1.09) |
| **São Paulo (reference)** | --- | --- |
| **Maranhão** | 1.15 (1.08, 1.22) | 1.00 (0.94, 1.07) |
| **Sergipe** | 1.25 (1.19, 1.30) | 1.01 (0.91, 1.13) |
| **Mato Grosso** | 1.41 (1.34, 1.48) | 1.03 (0.94, 1.12) |
| **Amazonas** | 1.25 (1.13, 1.38) | 1.03 (0.97, 1.08) |
| **Pernambuco** | 1.33 (1.23, 1.43) | 1.04 (0.99, 1.10) |
| **Tocantins** | 0.98 (0.79, 1.21) | 1.07 (0.84, 1.35) |
| **Amapá** | 0.88 (0.72, 1.07) | 1.08 (0.87, 1.33) |
| **Rondônia** | 1.17 (1.05, 1.31) | 1.14 (1.01, 1.28) |
| **Ceará** | 1.25 (1.19, 1.16) | 1.19 (1.13, 1.26) |
| **Santa Catarina** | 1.13 (1.06, 1.21) | 1.21 (1.12, 1.30) |
| **Rio De Janeiro** | 1.41 (1.36, 1.45) | 1.27 (1.23, 1.32) |
| **Rio Grande do Sul** | 1.78 (1.71, 1.85) | 1.33 (1.27, 1.39) |
| **Paraná** | 0.97 (0.91, 1.04) | 1.34 (1.25, 1.44) |
| **Mato Grosso do Sul** | 1.50 (1.38, 1.63) | 1.35 (1.23, 1.48) |
| **Roraima** | 1.24 (1.02, 1.50) | 1.67 (1.35, 2.06) |

Results ordered by adjusted odds-ratio.

**Table S2.** Adjusted odds ratios for unsuccessful treatment outcome stratified by year, 2015-2018.

| **Variables (reference)** | **Adjusted OR**<br>**(95% CI)**<br>**2015** | **Adjusted OR**<br>**(95% CI)**<br>**2016** | **Adjusted OR**<br>**(95% CI)**<br>**2017** | **Adjusted OR**<br>**(95% CI)**<br>**2018** |
| --- | --- | --- | --- | --- |
| **Age (25-34)** |  |  |  |  |
| 0-4 | 0.45 (0.35, 0.57) | 0.56 (0.45, 0.70) | 0.50 (0.40, 0.63) | 0.49 (0.39, 0.60) |
| 5-14 | 0.40 (0.32, 0.49) | 0.34 (0.27, 0.43) | 0.38 (0.31, 0.47) | 0.46 (0.38, 0.55) |
| 15-24 | 1.05 (0.97, 1.12) | 1.02 (0.95, 1.09) | 1.04 (0.98, 1.12) | 1.04 (0.98, 1.11) |
| 35-44 | 1.00 (0.94, 1.07) | 0.96 (0.89, 1.02) | 0.95 (0.90, 1.01) | 0.90 (0.84, 0.96) |
| 45-54 | 0.92 (0.85, 0.99) | 0.93 (0.86, 1.00) | 0.90 (0.96, 0.96) | 0.88 (0.82, 0.95) |
| 55-64 | 1.00 (0.93, 1.09) | 1.01 (0.93, 1.09) | 0.96 (0.89, 1.04) | 0.90 (0.84, 0.98) |
| 65-74 | 1.34 (1.22, 1.48) | 1.32 (1.20, 1.46) | 1.22 (1.11, 1.34) | 1.22 (1.12, 1.34) |
| 75-84 | 1.96 (1.73, 2.22) | 2.03 (1.80, 2.29) | 1.75 (1.56, 1.97) | 1.76 (1.56, 1.98) |
| 85+ | 2.53 (2.04, 3.13) | 3.67 (3.02, 4.47) | 2.84 (2.32, 3.47) | 2.91 (2.41, 3.52) |
| **Sex (Male)** |  |  |  |  |
| Female | 0.79 (0.75, 0.83) | 0.82 (0.78, 0.86) | 0.80 (0.76, 0.84) | 0.75 (0.72, 0.79) |
| **Race (White)** |  |  |  |  |
| Black | 1.24 (1.16, 1.33) | 1.29 (1.20, 1.38) | 1.26 (1.18, 1.35) | 1.29 (1.21, 1.38) |
| Yellow | 0.91 (0.69, 1.19) | 1.00 (0.72, 1.36) | 1.05 (0.82, 1.33) | 1.07 (0.84, 1.35) |
| Mixed | 1.12 (1.07, 1.18) | 1.17 (1.11, 1.24) | 1.10 (1.05, 1.15) | 1.11 (1.06, 1.17) |
| Indigenous | 1.08 (0.86, 1.34) | 0.85 (0.67, 1.07) | 0.92 (0.74, 1.15) | 1.03 (0.82, 1.28) |
| Other | 0.96 (0.88, 1.05) | 0.99 (0.91, 1.08) | 1.02 (0.93, 1.11) | 1.03 (0.94, 1.12) |
| **Education (Complete high school)** |  |  |  |  |
| No education | 1.90 (1.67, 2.17) | 1.29 (1.57, 2.04) | 2.05 (1.10, 1.33) | 1.95 (1.73, 2.20) |
| Incomplete 1-4<sup>th</sup> grade | 1.76 (1.59, 1.96) | 1.71 (1.55, 1.90) | 1.77 (0.43, 0.59) | 1.74 (1.58, 1.92) |
| Complete 1-4<sup>th</sup> grade | 1.71 (1.56, 1.89) | 1.65 (1.50, 1.81) | 1.82 (0.54, 0.64) | 1.72 (1.58, 1.88) |
| Complete 5-8<sup>th</sup> grade | 1.34 (1.21, 1.48) | 1.33 (1.20, 1.46) | 1.47 (0.34, 0.47) | 1.45 (1.33, 1.59) |
| Any higher education | 0.77 (0.67, 0.88) | 0.82 (0.72, 0.94) | 0.77 (1.09, 1.21) | 0.74 (0.65, 0.83) |
| Other | 1.91 (1.74, 2.10) | 1.82 (1.66, 2.00) | 1.84 (1.68, 2.01) | 1.92 (1.77, 2.09) |
| **Diabetes (no)** |  |  |  |  |
| Yes | 0.97 (0.89, 1.05) | 0.98 (0.90, 1.07) | 0.89 (0.82, 0.96) | 0.91 (0.84, 0.98) |
| Other | 0.94 (0.83, 1.08) | 0.96 (0.84, 1.10) | 0.86 (0.75, 0.98) | 0.95 (0.83, 1.08) |
| **HIV (negative)** |  |  |  |  |
| Yes | 3.22 (3.02, 3.45) | 2.97 (2.78, 3.18) | 2.68 (2.51, 2.86) | 2.60 (2.43, 2.77) |
| Other | 1.99 (1.89, 2.10) | 1.89 (1.79, 1.99) | 1.77 (1.68, 1.87) | 1.76 (1.67, 1.86) |
| **Smoking (no)** |  |  |  |  |
| Yes | 1.21 (1.13, 1.28) | 1.17 (1.10, 1.24) | 1.17 (1.11, 1.24) | 1.19 (1.13, 1.26) |
| Other | 1.00 (0.87, 1.16) | 0.89 (0.89, 1.20) | 1.29 (1.12, 1.49) | 1.29 (1.11, 1.50) |
| **Alcohol (no)** |  |  |  |  |
| Yes | 1.47 (1.38, 1.56) | 1.46 (1.37, 1.55) | 1.42 (1.34, 1.50) | 1.35 (1.27, 1.43) |
| Other | 0.98 (0.85, 1.13) | 1.10 (0.95, 1.27) | 1.00 (0.87, 1.16) | 1.01 (0.87, 1.17) |
| **Illicit drug use (no)** |  |  |  |  |
| Yes | 1.83 (1.70, 1.97) | 2.06 (1.91, 2.21) | 1.94 (1.82, 2.07) | 1.97 (1.85, 2.10) |
| Other | 1.17 (1.03, 1.34) | 1.23 (1.07, 1.42) | 1.16 (1.01, 1.34) | 1.22 (1.06, 1.40) |
| **Incarcerated (no)** |  |  |  |  |
| Yes | 0.55 (0.50, 0.61) | 0.51 (0.46, 0.56) | 0.54 (0.49, 0.58) | 0.49 (0.45, 0.53) |
| Other | 1.01 (0.82, 1.25) | 1.13 (0.87, 1.47) | 1.13 (0.89, 1.43) | 1.00 (0.78, 1.28) |
| **Homeless (no)** |  |  |  |  |
| Yes | 2.90 (2.57, 3.27) | 3.19 (2.83, 3.60) | 3.49 (3.11, 3.92) | 3.12 (2.79, 3.48) |
| Other | 0.97 (0.78, 1.21) | 0.88 (0.67, 1.17) | 0.86 (0.64, 1.16) | 0.84 (0.61, 1.14) |
| **Immigrants (no)** |  |  |  |  |
| Yes | 1.52 (1.11, 2.06) | 1.26 (0.96, 1.62) | 1.15 (0.87, 1.51) | 1.48 (1.18, 1.85) |
| Other | 1.04 (0.94, 1.15) | 0.94 (0.79, 1.11) | 0.98 (0.74, 1.28) | 1.07 (0.81, 1.42) |
| **Health unit (primary care)** |  |  |  |  |
| Secondary care | 1.21 (1.15, 1.27) | 1.14 (1.09, 1.21) | 1.17 (1.11, 1.23) | 1.24 (1.18, 1.30) |
| Tertiary care | 2.15 (2.02, 2.29) | 2.09 (1.96, 2.22) | 2.14 (2.02, 2.27) | 2.20 (2.08, 2.33) |
| Other | 0.98 (0.87, 1.11) | 1.07 (0.95, 1.21) | 0.96 (0.85, 1.08) | 1.06 (0.94, 1.19) |
| **DOT (yes)** |  |  |  |  |
| No | 2.59 (2.45, 2.75) | 2.30 (2.17, 2.43) | 2.40 (2.27, 2.53) | 2.38 (2.27, 2.51) |
| Other | 3.56 (3.35, 3.78) | 3.13 (2.95, 3.32) | 3.26 (3.07, 3.45) | 3.09 (2.93, 3.27) |
| **Bacteriological test (positive)** |  |  |  |  |
| Negative | 1.10 (1.03, 1.17) | 1.15 (1.08, 1.22) | 1.15 (1.09, 1.23) | 1.17 (1.10, 1.24) |
| Not determined | 1.32 (1.23, 1.40) | 1.37 (1.29, 1.46) | 1.30 (1.22, 1.38) | 1.33 (1.25, 1.41) |
| **Chest X-ray (suggestive of TB)** |  |  |  |  |
| Normal | 1.00 (0.91, 1.10) | 1.02 (0.93, 1.13) | 1.03 (0.94, 1.13) | 1.00 (0.92, 1.10) |
| Not performed | 1.02 (0.97, 1.08) | 1.05 (0.99, 1.11) | 1.03 (0.97, 1.08) | 1.07 (1.01, 1.12) |
| **Type of TB (pulmonary)** |  |  |  |  |
| Extrapulmonary | 1.07 (0.95, 1.20) | 1.01 (0.90, 1.14) | 1.04 (0.93, 1.17) | 1.01 (0.90, 1.13) |
| Both | 0.70 (0.65, 0.76) | 0.73 (0.68, 0.79) | 0.70 (0.65, 0.75) | 0.68 (0.63, 0.73) |

**Table S3.** Adjusted risk ratios for categorical treatment outcome, 2015-2018.

| Variables (reference) | Adjusted RR (95% CI) |  |
| --- | --- | --- |
|  | Loss to follow-up | Death |
| **Age (25-34)** |  |  |
| 0-4 | 0.50 (0.44, 0.58) | 0.71 (0.59, 0.84) |
| 5-14 | 0.47 (0.42, 0.53) | 0.36 (0.29, 0.45) |
| 15-24 | 1.11 (1.07, 1.15) | 0.74 (0.69, 0.80) |
| 35-44 | 0.86 (0.83, 0.89) | 1.34 (1.27, 1.42) |
| 45-54 | 0.62 (0.60, 0.65) | 1.95 (1.84, 2.06) |
| 55-64 | 0.45 (0.43, 0.48) | 2.94 (2.77, 3.12) |
| 65-74 | 0.39 (0.36, 0.42) | 4.66 (4.37, 4.98) |
| 75-84 | 0.55 (0.41, 0.51) | 7.42 (6.87, 8.01) |
| 85+ | 0.80 (0.45, 0.67) | 11.83 (10.54, 13.28) |
| **Sex (Male)** |  |  |
| Female | 0.80 (0.77, 0.82) | 0.80 (0.77, 0.82) |
| **Race (White)** |  |  |
| Black | 1.40 (1.34, 1.46) | 1.08 (1.03, 1.14) |
| Yellow | 1.20 (1.03, 1.40) | 0.86 (0.70, 1.06) |
| Mixed | 1.18 (1.15, 1.22) | 1.04 (1.01, 1.08) |
| Indigenous | 0.99 (0.86, 1.14) | 0.91 (0.77, 1.07) |
| Other | 1.10 (1.04, 1.16) | 0.87 (0.81, 0.93) |
| **Education (Complete high school)** |  |  |
| No education | 1.95 (1.80, 2.12) | 2.03 (1.86, 2.22) |
| Incomplete 1-4<sup>th</sup> grade | 1.78 (1.67, 1.89) | 1.75 (1.62, 1.89) |
| Complete 1-4<sup>th</sup> grade | 1.82 (1.72, 1.92) | 1.54 (1.43, 1.65) |
| Complete 5-8<sup>th</sup> grade | 1.47 (1.39, 1.55) | 1.25 (1.16, 1.35) |
| Any higher education | 0.78 (0.72, 0.85) | 0.78 (0.70, 0.86) |
| Other | 1.74 (1.65, 1.84) | 2.07 (1.93, 2.22) |
| **Diabetes (no)** |  |  |
| Yes | 0.81 (0.76, 0.86) | 1.10 (1.05, 1.16) |
| Other | 0.98 (0.91, 1.06) | 0.90 (0.82, 0.99) |
| **HIV (negative)** |  |  |
| Yes | 1.92 (1.84, 2.00) | 5.09 (4.87, 5.32) |
| Other | 1.91 (1.85, 1.97) | 1.79 (1.72, 1.86) |
| **Smoking (no)** |  |  |
| Yes | 1.22 (1.18, 1.26) | 1.11 (1.07, 1.16) |
| Other | 1.05 (0.96, 1.15) | 1.26 (1.14, 1.39) |
| **Alcohol (no)** |  |  |
| Yes | 1.36 (1.31, 1.41) | 1.59 (1.52, 1.66) |
| Other | 1.01 (0.92, 1.10) | 1.09 (0.98, 1.20) |
| **Illicit drug use (no)** |  |  |
| Yes | 2.19 (2.11, 2.27) | 1.29 (1.21, 1.37) |
| Other | 1.18 (1.09, 1.29) | 1.19 (1.08, 1.31) |
| **Incarcerated (no)** |  |  |
| Yes | 0.54 (0.52, 0.57) | 0.44 (0.40, 0.48) |
| Other | 0.95 (0.83, 1.09) | 1.04 (0.86, 1.24) |
| **Homeless (no)** |  |  |
| Yes | 3.82 (3.59, 4.06) | 2.11 (1.93, 2.31) |
| Other | 0.90 (0.78, 1.04) | 0.88 (0.73, 1.07) |
| **Immigrants (no)** |  |  |
| Yes | 1.43 (1.24, 1.66) | 1.19 (0.96, 1.48) |
| Other | 1.11 (1.02, 1.22) | 1.00 (0.89, 1.13) |
| **Health unit (primary care)** |  |  |
| Secondary care | 1.01 (0.98, 1.04) | 1.74 (1.67, 1.82) |
| Tertiary care | 1.24 (1.19, 1.29) | 4.46 (4.28, 4.66) |
| Other | 0.81 (0.76, 0.88) | 1.60 (1.46, 1.75) |
| **DOT (yes)** |  |  |
| No | 3.06 (2.96, 3.16) | 1.56 (1.49, 1.63) |
| Other | 3.43 (3.31, 3.56) | 2.88 (2.76, 3.00) |
| **Bacteriological test (positive)** |  |  |
| Negative | 1.00 (0.96, 1.04) | 1.43 (1.37, 1.50) |
| Not determined | 1.16 (1.12, 1.21) | 1.68 (1.61, 1.75) |
| **Chest X-ray (suggestive with TB)** |  |  |
| Normal | 0.94 (0.89, 1.00) | 1.05 (0.98, 1.12) |
| Not performed | 1.09 (1.06, 1.13) | 0.91 (0.87, 0.95) |
| **Type of TB (pulmonary)** |  |  |
| Extrapulmonary | 0.81 (0.75, 0.87) | 1.38 (1.28, 1.48) |
| Both | 0.66 (0.63, 0.79) | 0.76 (0.72, 0.80) |

